# Mapping the retail food environment at high resolution across Europe

**DOI:** 10.64898/2026.07.29.26359016

**Authors:** Mohan Raju JS, Kamille Almer Bernsdorf, Alfred Wagtendonk, Nishit Patel, Julia Díez, Jet D.S. van de Geest, Roberto Valiente, Anna Bartoskova, Thomas Burgoine, Jeroen Lakerveld

**Affiliations:** Department of Epidemiology & Data Science, Amsterdam Public Health research institute, Amsterdam UMC, the Netherlands; Amsterdam Public Health Research Institute, 1105 AZ, Amsterdam, the Netherlands; Upstream Team, www.upstreamteam.nl, Amsterdam UMC, VU University Amsterdam, De Boelelaan 1117, 1081 HV, Amsterdam, Netherlands; Department for Prevention, Steno Diabetes Center Copenhagen, Denmark; Public Health and Epidemiology Research Group, School of Medicine and Health Sciences, Universidad de Alcalá, Alcalá de Henares, Madrid, Spain; Faculty of Science, Masaryk University, Brno, Czech Republic; IMS Epidemiology, University of Cambridge School of Clinical Medicine, Cambridge, United Kingdom

**Keywords:** Retail food environment, Food outlet classification, Google Maps, Validation study, Neighbourhood co-exposures, Europe

## Abstract

**Aim:** Neighbourhood food outlet availability influences dietary behaviours and nutrition related health outcomes. However, food environment research across Europe remains limited by inconsistent spatial data, heterogenous food outlet classifications, and the lack of publicly available high-resolution retail food data. We developed and evaluated a scalable framework to acquire, classify, validate, and analyse food-related points of interest (POIs) from Google Maps across 39 countries in Europe and Turkey. We also developed a method for characterizing the joint co-occurrence of multiple outlet types as neighbourhood-level food environment exposures.

**Methods:** We harmonized ∼3.26 million food-related POIs. Outlets were classified into five main categories (cafés and bakeries, restaurants, fast-food and snack outlets, groceries and food retail, and bars and pubs) and 25 subcategories. We assessed data quality against official register**s** in five regions (the Netherlands, United Kingdom, Denmark, Madrid (Spain), and Brno (Czech Republic)), testing agreement in positional accuracy, completeness, spatial clustering (Nearest Neighbour Index), and spatial density (kernel density estimation (KDE)). To enable cross-country comparisons, POIs were aggregated to national and city scales, with food outlet density calculated as outlets per 1,000 residents and standardised using z-scores. To characterize neighbourhood-level co-exposures, we applied Principal Component Analysis (PCA) followed by Latent Class Analysis (LCA) to z-standardized outlet densities across 500 m hexagonal grid cells in eight major European cities.

**Results:** Google Maps data showed high positional accuracy, with 91% of outlets located within 30m of registry records. Completeness varied by region, while KDE comparisons showed moderate-to-strong spatial agreement with city-specific variation. PCA identified two components explaining 68.4% of the variance (PC1: 51.3%, PC2: 17.1%), with PC2 differing between café/bar and fast-food/supermarket densities. LCA identified four neighbourhood classes: low-access, moderate-mixed, dining-out, and high-density-mixed.

**Conclusion:** Our harmonized high-resolution framework supports cross-national monitoring of retail food environments and spatial epidemiological research across European settings.

## Introduction

Unhealthy dietary behaviours are key contributors to obesity, type 2 diabetes, cardiovascular disease, and several cancers (*World Health Organization*, 2023). Such behaviours are partly driven by the wider food environment, which includes the retail food environment. The retail food environment refers to the places where people purchase and eat food and is increasingly studied as one of several spatially co-occurring urban exposures that may collectively shape population health. For example, greater fast-food outlet density has been associated with greater obesity risk and cardiovascular disease prevalence, while greater supermarket accessibility has been linked to lower obesity risk (Meijer et al., 2023; Pineda et al., 2024). However, findings across studies remain inconsistent, partly owing to methodological differences in exposure assessment, including variation in neighbourhoods, data sources, food outlet classification, exposure metrics, and temporal alignment (Pinho et al., 2019a; Pinho et al., 2019b).

A persistent methodological challenge is the absence of a standardized food outlet classification system that can be applied consistently across countries and regions. Food retail landscapes, outlet naming conventions, and the functional roles of outlets vary substantially across and within countries. Existing studies have used a range of audit tools and classification systems, often tailored to specific national or local contexts, limiting cross-national comparability (Caspi et al., 2012; Pinho et al., 2019a). These differences highlight the need for a standardized but context-sensitive classification approach that supports consistent -European food environment mapping while accommodating regional variation in food retail structures.

Geographic Information Systems (GIS) and spatial analysis of secondary data are now dominant approaches for characterising physical access to food outlets (Lytle and Sokol, 2017). Secondary data sources vary substantially in accuracy and completeness across databases and settings. While field audits remain the gold standard, administrative records, commercial databases, and virtual audits can capture the presence of food outlets with good to excellent accuracy in many contexts (Bader *et al*., 2010; Fleischhacker *et al*., 2013; Bethlehem *et al*., 2014). Nonetheless, assessing the validity of secondary food outlet data remains uncommon and unmeasured variation in data quality could introduce bias in exposure assessment, particularly in cross-national studies (Lytle and Sokol, 2017).

Google Maps has emerged as a promising data source for food environment mapping, providing geocoded information on outlet type, location, and operational status across many countries (Präger, 2024). Researchers can compile geocoded food retail data at multiple geographic scales through the Google Maps Application Programming Interface (API, 2024). Previous validation studies from other continents have shown that Google-based food outlet data can perform well in different contexts, although validity varies by outlet type, data source, and local context. For example, in Brazil, sensitivity ranged from 66.3% to 100% with overall accuracy of 99.1% (de Menezes *et al*., 2021), while in Mexico, agreement with official records was moderate but systematically weaker in lower-income neighbourhoods (Ramírez-Toscano *et al*., 2024).Systematic, harmonized mapping of retail food outlets across Europe is therefore essential. Food environments differ widely between countries in Europe owing to differences in retail systems, urban form, planning contexts, food cultures and regulatory environments. Yet research across Europe remains constrained by spatial variability in data availability, limited access to harmonised food retail data at high spatial resolution, and inconsistent methods for validating food outlet data across geographical regions (Wilkins *et al*., 2017). Harmonised maps can support comparative analyses of food outlet access, reveal geographic and socioeconomic inequalities, and provide a validated basis for linking food environments to dietary behaviours and health outcomes. A key scientific gap is the absence of a validated, cross-national methodological framework that integrates data acquisition, classification, spatial validation, and multiscale exposure construction consistently across European settings.

Moreover, most food environment research characterises exposures using single outlet types, such as supermarkets or fast-food outlets, rather than the combined food retail landscape that residents encounter in everyday life. Food outlet types co-occur spatially, with neighbourhoods with many fast-food outlets tending also to contain many other types of food outlets e.g. restaurants, cafés, supermarkets. These co-occurring exposures interact with broader urban environmental features in ways that univariate metrics cannot capture (Song *et al*., 2022; de Hoogh *et al*., 2025). Characterising the joint distribution of multiple food outlet types as simultaneous neighbourhood-level exposures requires multivariate spatial methods, yet this approach remains underdeveloped in European food environment research. Such methods are also essential for integrating food environment data into broader multi-exposure urban health frameworks, where interacting, cumulative and spatially co-occurring neighbourhood exposures are increasingly recognised as central to understanding population health (Wilkins *et al*., 2017; Song *et al*., 2022).

This methodological study therefore had three aims: (1) to acquire large-scale food outlet data across Europe using Google Maps and develop a standardised, cross-nationally applicable food outlet classification system; (2) to assess the positional accuracy, completeness, and geographic consistency of Google-derived points of interest against official food retail registries in five European regions; and (3) to characterise the distribution, density, and co-occurring spatial patterns of food outlet types across European countries and major cities using spatial and multivariate analytical methods.

## Methods

The retail food environment mapping framework presented here was developed within the European Commission-funded project OBCT project (Obesity: Biological, socioCultural, and environmental risk Trajectories) (Lam *et al*., 2025).

### 1. Study area

The study area was 39 countries across a broad European study region or nearly 6 million km². It included the 27 European Union (EU) member states (Eurostat, 2025), the four European Free Trade Association (EFTA) countries (Iceland, Liechtenstein, Norway, and Switzerland), six Western Balkan countries (Albania, Bosnia and Herzegovina, Kosovo, Montenegro, North Macedonia, and Serbia), as well as the United Kingdom and Turkey. Turkey was included as its foodscape has not yet been studied, to reflect the broader European study region and to capture wider geographic variation in food environment analysis.

Administrative boundaries were obtained from Eurostat geospatial boundary datasets. All spatial data were harmonised and projected to ETRS89 / LAEA Europe coordinate reference system, which is commonly used for pan-European spatial analyses and ensures compatibility with standardised EU-wide data sources (EPSG:3035, 2021). Geoprocessing, spatial aggregation, and validation analyses were conducted using this common projection to minimize inconsistencies arising from cross-country differences in coordinate systems and boundary definitions.

### 2. Food outlet data acquisition

We employed a systematic keyword-based query approach using the Google Maps Places Application Programming Interface (API), to retrieve food-related points of interest (POIs) across all study regions. Data were collected in September 2025.

The search strategy was developed by cross-referencing retail categories from the Locatus commercial database (Locatus, 2024), a field-audited and epidemiologically validated source of retail outlet classifications in the Netherlands (Canalia *et al*., 2020), with the keyword framework validated by Bernsdorf *et al*. (2022) for European retail food environment assessment, and with the official Google Maps Places API place types documentation (Google Places API, 2026). This cross-referencing yielded a harmonised set of search terms and place types covering diverse food outlet types including restaurants, supermarkets, convenience stores, cafés, bakeries, fast-food outlets, bars, and other food-related retail locations.

To minimise misclassification and improve data quality, we performed an iterative refinement process. We conducted test queries across selected representative geographic areas and reviewed the results manually to identify food outlets and exclude non-food outlets that were incorrectly retrieved (Figure 1). This process resulted in a final, targeted set of 14 standardised Google place types, selected to maximise coverage of relevant food outlets across all study regions while limiting the inclusion of unrelated POIs. Provided in supplementary file: table-S2.

**Figure 1.**
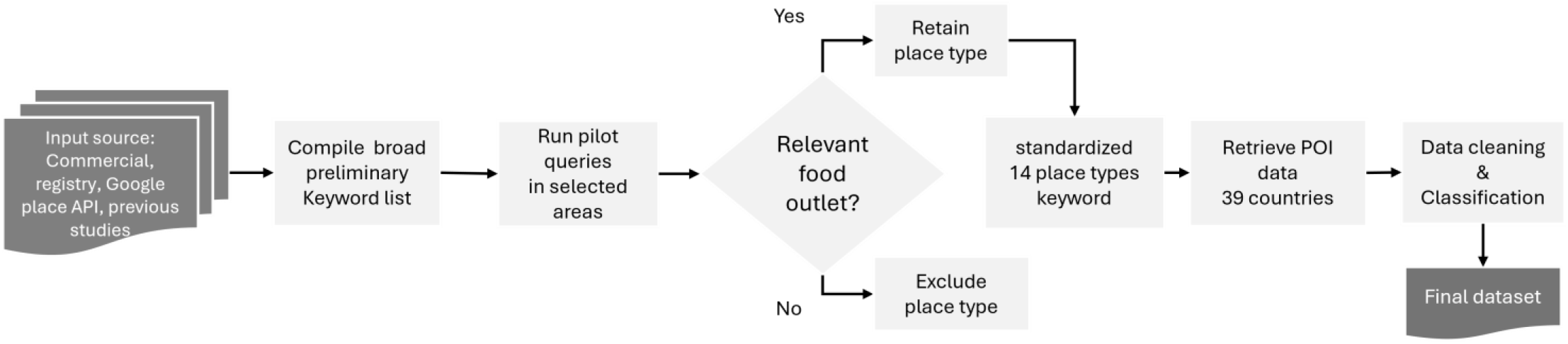
Workflow for acquiring, refining, and cleaning Google Maps food-related points of interest across the European study region.

For every identified (POI), we obtained the following details in the form of structured metadata: outlet name, type, latitude, longitude, address, and country code. Duplicate records, defined as entries sharing identical values across all five metadata fields simultaneously, were identified and removed to ensure that each food outlet was represented only once in the final dataset.

### 3. Food outlet classification

We developed a harmonised food outlet classification system to ensure consistent categorisation across diverse European retail food environments. The system drew on the 22-category classification tool developed by Lake et al. (2010), the European-specific food outlet categories validated by Bernsdorf et al.(2024) and the retail categories used in the Locatus commercial database (Locatus, 2024).

The system was further refined by an inductive classification process based on 600 Google place- type and business-category variants retrieved during the data acquisition stage, such as *mexican_restaurant*, *supermarket*, *grocery_stor*e. This process ensured that regional naming variations, outlet subtypes, and food-related categories not fully captured by existing tools were systematically assigned to harmonised categories. This process yielded a final classification of five main categories and 25 subcategories (Table 1). The five main categories were used in the present analyses, while the 25 subcategories were retained to preserve more detailed outlet information and support future analyses. Classification was applied through the following three-stage harmonisation process, described below:

**Table 1:** European-wide harmonized classification of food outlet categories and subcategories.

| Main category | 1st sub-category | 2nd sub-category |
| --- | --- | --- |
| Sweets & cafés | Cafés |  |
|  | Sweet bakeries /dessert shops |  |
|  | Candy stores |  |
| Restaurants | Asian |  |
|  | Bistros |  |
|  | Meat/grill/barbecue |  |
|  | Mediterranean |  |
|  | Middle Eastern |  |
|  | Regional/country cuisine |  |
|  | Seafood/fish |  |
|  | Vegetarian/vegan |  |
|  | Other/non-cuisine specific |  |
| Fast-food & snack outlets | Snackbars focused on (deep-)fried food/Fast food chains |  |
|  | Quick-meal/snack outlets |  |
| Groceries & food retail | Local/specialty food retailers | Meat & poultry retailers |
|  |  | Dairy retailers |
|  |  | Fish retailers |
|  |  | Bakery shop (bread-focused) |
|  |  | Other specialty food retailers |
|  | Grocery stores/green groceries |  |
|  | Convenience stores |  |
|  | Franchise supermarkets |  |
|  | Off-licenses/liquor stores |  |
| Bars & pubs | Bars/Pubs |  |

#### 3.1 Direct mapping of place types

First, standardised Google Places types and business-category labels were directly mapped onto one of the five main food outlet categories. For example, place types such as ‘*sushi_restaurant*’ and ‘*indian_restaurant*’ were grouped under the broader category of Restaurants. Each outlet was also assigned to a more specific subcategory to retain additional classification detail.

#### 3.2 Harmonisation of regional variants

Second, regional and linguistic variation in Google-assigned business labels was harmonised by grouping redundant, overlapping, or locally specific place types into standardised subcategories. This step reduced inconsistencies arising from differences in naming conventions, language, and retail structures across study regions.

#### 3.3 Manual review of ambiguous classifications

Third, outlets with ambiguous, vague, or unspecified classifications were flagged for manual review. These included outlets with broad food-related labels, unclear business names, or categories that could refer to either food and non-food establishments. Manual review was used to determine whether each outlet was relevant to the retail food environment system and to assign it to the most appropriate main category and subcategory. Only outlets considered relevant to the retail food environment were retained in the final classified dataset.

### 4. Food outlet validation framework

To assess the validity of the Google POI data, we applied a multi-region validation framework across five study regions. We selected these regions based on the availability of suitable governmental or commercial registry data serving as ‘silver’-standard reference sources. Registry datasets were available at different geographic scales: country-level data were obtained for the Netherlands, Denmark, and the United Kingdom, while city-level data were available for the municipality of Brno and the municipality of Madrid. Provided in supplementary file: **table S3**.

We assessed Google POI validity using four complementary metrics: positional accuracy, completeness, spatial clustering, and spatial density agreement(Sections 4.1-4.3) were conducted at the scale at which registry data were available; either country or city level. Spatial density analysis using KDE (Section 4.4) was conducted specifically at city scale across all five regions, as rural and peri-urban outlet density is sparse enough that density surfaces become highly sensitive to individual point locations, and urban contexts are where population exposure to food outlets is most consequential for health outcomes (Apparicio et al., 2008; Kestens et al., 2012) (Table 2).

**Table 2.** Components, methods, and rational of the validation framework for Google-derived food outlet POIs data against registry datasets.

| Validation component | Operational method | Rationale |
| --- | --- | --- |
| Positional accuracy | Point-to-point spatial matching | To assess how closely the coordinates of Google-derived POIs aligned with corresponding registry outlet locations. |
| Completeness | Bidirectional correspondence matching | To assess the extent to which outlets recorded in each dataset were represented in the other. |
| Spatial clustering | Nearest Neighbour Index (NNI) | To compare whether Google POIs and registry outlets exhibited similar degrees of spatial clustering or dispersion. |
| Spatial density agreement | Kernel Density Estimation (KDE) | To examine whether Google POIs and registry datasets showed similar continuous spatial intensity patterns in urban areas. |

#### 4.1 Positional accuracy

Positional accuracy was assessed by measuring the Euclidean distance between matched outlet locations from the Google POI and registry datasets within each validation region. Outlet pairs were considered matched when they represented the same or equivalent food outlet type and fell within the specified spatial matching threshold. Directional displacement (easting and northing coordinates) was also calculated to assess whether Google POI systematically shifts outlet locations relative to registry data, which would indicate geocoding bias (Lih Wei Yeow et al., 2021).

We applied distance thresholds of 10, 20, and 30 m to evaluate positional accuracy at increasingly permissive tolerance levels. Outcome metrics included the number of matched outlet pairs, median distance error, standard deviation of the distance error, median easting and northing displacement, and the proportion of matched outlets falling within each distance thresholds.

#### 4.2 Completeness

To evaluate completeness beyond simple outlet counts, we implemented a bidirectional, correspondence-based validation approach. Outlets in each dataset were assessed according to whether a corresponding outlet was present in the other dataset, based on spatial proximity and food outlet type. Two complementary proportions were calculated for each validation region: (1) the proportion of registry outlets matched to a Google POI data, and (2) the proportion of Google POIs matched to a registry outlet.

This bidirectional approach distinguishes between records missing from one dataset (omission) and excess or potentially spurious records in the other (commission), and is preferable to aggregate count comparisons for this reason (Lih Wei Yeow et al., 2021). Evaluating completeness in both directions provides a more informative assessment of relative cross-dataset coverage than total feature counts alone.

#### 4.3 Spatial clustering

We calculated the Nearest Neighbour Index (NNI) separately for Google POIs and registry POIs within each validation region to assess whether food outlets showed clustered, random, or dispersed spatial patterns (Clarks and Evans, 1954). The NNI compares the observed average nearest-neighbour distance to the expected average distance under complete spatial randomness. Values below 1.0 indicate clustering, values above 1.0 indicate dispersion, and values near 1.0 indicate a pattern consistent with spatial randomness. Similarity in NNI values between Google POI and registry datasets was interpreted as evidence of comparable spatial patterning of food outlets.

#### 4.4 Spatial density agreement

Kernel Density Estimation (KDE) was applied to both Google POI and registry datasets to generate continuous food outlet density surfaces depicting food outlet concentration across each validation city, using a 500 m kernel radius. KDE outputs were compared visually to assess spatial consistency, with aligned high-density values indicating agreement in concentration patterns and divergence indicating potential coverage gaps or over-representation (Kestens *et al*., 2012). Pearson correlations between Google POI and registry KDE surfaces were calculated to quantify overall spatial density agreement. The KDE maps served primarily as a visual and exploratory complement to the quantitative metrics, enabling identification of systematic spatial patterns of agreement or disparity.

### 5. Two-scale spatial analysis of food outlet distribution across Europe

To quantify food environment distribution across Europe, we analysed food outlet distribution at two distinct spatial scales: countries (n=39) and 500m hexagonal neighborhoods for the eight most populous European cities (Amsterdam, Berlin, Budapest, Copenhagen, Lisbon, Madrid, Paris, Warsaw). This two-scale approach supports cross-country comparisons of food system structure at the national level, and detailed characterisation of neighbourhood-level food environment variation and co-exposure patterns at the city level.

#### 5.1 Share of food outlet categories

To examine the compositional structure of food environments across European countries, POIs were aggregated at the national level. The proportional share of each of the five main categories (see Table 1) was calculated relative to the total number of food outlets per country, enabling cross-national comparison of food environment composition.

#### 5.2 Food outlet density per 1,000 population

Food outlet density was calculated at the country level as:

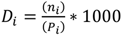

where *Di* is the food outlet density for country *i*, *n_i_* is the total number of food outlets, and *P_i_* is the total resident population, based on Eurostat population data (2024).

#### 5.3 Standardized food outlet density

To enable cross-country comparisons that account for structural differences in national food environments, outlet densities were standardized using z-scores calculated separately within each main outlet category across all countries. The z-score for each country was computed as:

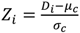

where *Z_I_* is the standardized density for country *i*, *D_i_* is the raw outlet density per 1,000 population for country *i*, *μ_c_* is the mean outlet density across all countries for category c, and *σ_c_* is the corresponding population standard deviation. Positive z-scores indicate countries with above-average outlet density relative to the European mean, while negative values indicate below-average density.

#### 5.4 Characterizing joint food environment exposures

To examine how food outlet co-occur at the neighbourhood level, a two-stage multivariate analysis was applied to population-standardised, z-score normalised outlet densities at the 500 m hexagonal grid level across the eight cities listed above. Hexagons with fewer than 50 residents were excluded to avoid unstable density estimates, and densities were capped at the 99th percentile to limit the influence of extreme values.

Principal Component Analysis (PCA) was first applied to identify the main patterns of co-variation among the five food outlet categories, with components interpreted based on loadings exceeding an absolute value of 0.40, indicating a meaningful contribution of that outlet category to the component.

Latent Class Analysis (LCA) was then applied to group hexagons into discrete neighbourhood food environment classes based on the simultaneous distribution of all five food outlet densities. Gaussian Mixture Modelling (GMM) was used as the computational implementation of LCA, as input variables are continuous z-scores rather than categorical indicators. Under this framework, each hexagon i is assigned to its most probable class k based on the posterior probability:

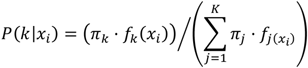

where:

- *π_k_* is the mixing proportion of class k
- *P(k∣xi) =posterior probability that neighbourhood i belongs to class k*
- *f_k_*(*x_i_*) is the multivariate Gaussian density of hexagon i under class k
- 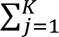 sum over all K latent classes

Models were estimated for two through seven classes, with the optimal solution selected based on the Bayesian Information Criterion (BIC), a statistical measure of model fit that penalises unnecessary complexity, alongside consideration of class size and interpretability. Each hexagon was assigned to the class with the highest posterior probability, and classes were characterised by their mean z-score per outlet category (Hobbs et al., 2018; DeWeese et al., 2018).

## Results

We acquired a total of 3,259,840 georeferenced food outlet POIs across 39 countries in the European study region. After cleaning and deduplication, POIs were classified into five main food outlet categories: restaurants (41.1%), groceries and food retail (31.5%), sweets and cafés (11.0%), fast-food and snack outlets (10.8%), and bars/pubs (2.7%).

### Validation

Table 3 presents positional accuracy metrics for matched Google-derived POIs and registry records across all five validation regions. Positional accuracy was high across all regions, with 100% of matched POIs falling within 30 m of their corresponding registry locations. The Netherlands demonstrated the strongest performance (median = 7.34 m; 65.5% within 10 m), while the United Kingdom showed the widest error spread (median = 9.56 m; 52.2% within 10 m). Denmark, Madrid, and Brno occupied an intermediate range, with median errors between 7.93 m and 9.23 m and between 54% and 59% of POIs within 10 m. The distribution of Euclidean positional displacement across outlet categories and regions is shown in supplementary file: Figure S1.

**Table 3.** Positional accuracy of matched Google-derived food outlet POIs compared with registry data, by validation region.

| Region | Matched pairs (n) | Median distance error (m) | SD (m) | % ≤ 10 m | % ≤ 20 m | % ≤ 30 m |
| --- | --- | --- | --- | --- | --- | --- |
| Netherlands | 52,798 | 7.34 | 6.48 | 65.5% | 91.5% | 100% |
| United Kingdom | 203,376 | 9.56 | 7.83 | 52.2% | 82.5% | 100% |
| Denmark | 11,332 | 7.93 | 7.58 | 59.3% | 85.1% | 100% |
| Madrid | 30,629 | 9.23 | 7.91 | 53.6% | 82.3% | 100% |
| Brno | 1,670 | 8.74 | 7.51 | 55.9% | 84.3% | 100% |

Figure 2 presents bidirectional completeness between Google-derived POIs and registry datasets across the five validation regions. Relative completeness varied substantially by region, with registry-to-Google correspondence ranging from 57.4% in Denmark to 92.2% in the Netherlands, while Google-to-registry correspondence ranging from 65.6% in Denmark to 94.0% in Madrid. Brno demonstrated the most balanced bidirectional coverage, with 83.8% of registry outlets matched to Google POIs and 78.3% of Google POIs matched to registry records. The United Kingdom and Denmark showed the weakest alignment in both directions, partly reflecting registry limitations: the UK registry did not include cafés and bakeries, and the Danish Smiley registry lacked coordinates for a substantial proportion of outlets, rendering them ineligible for spatial matching.

**Figure 2.**
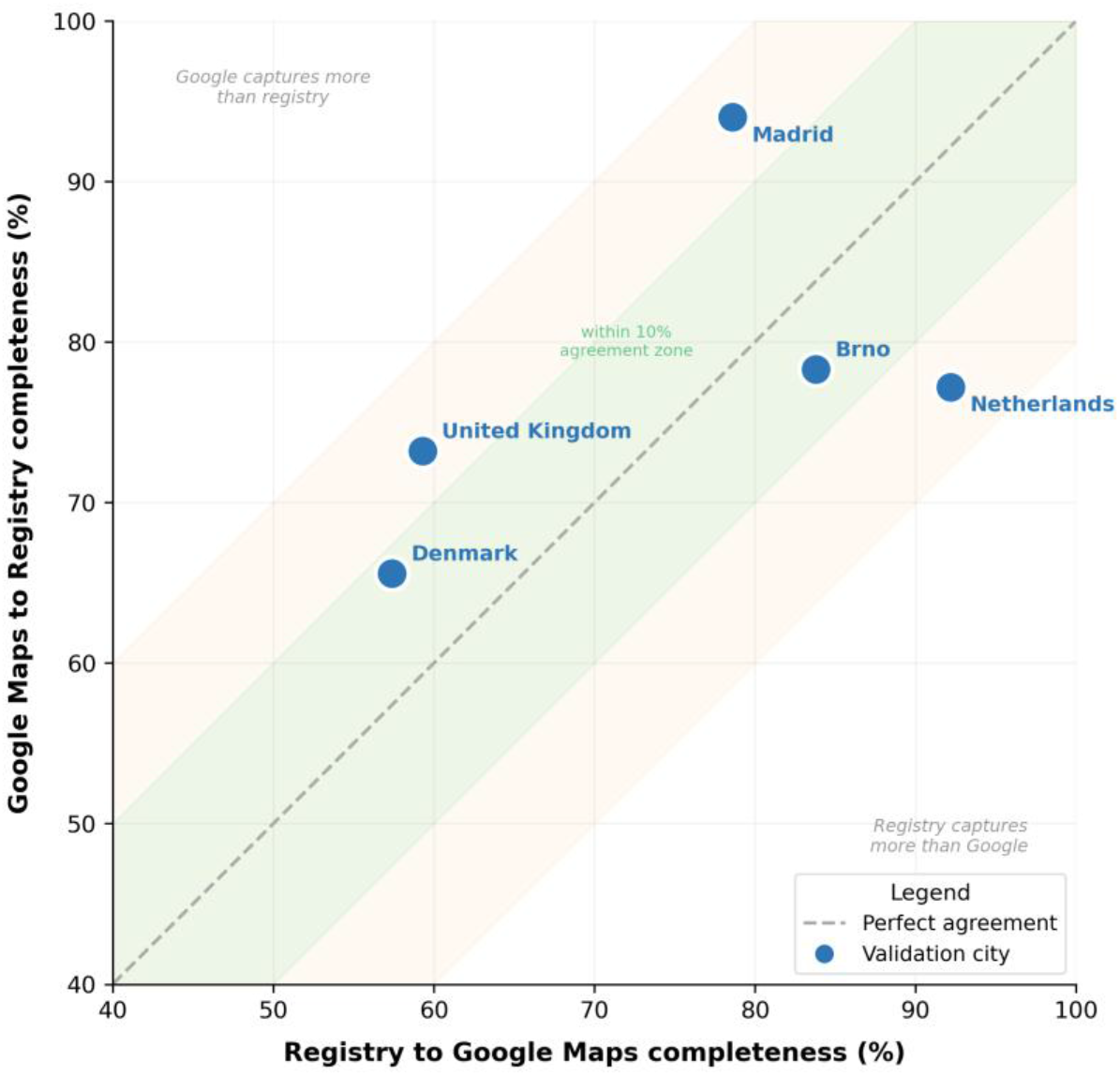
Bidirectional relative completeness between Google-derived food outlet POIs and registry datasets across five validation regions

Figure 3 presents NNI-based clustering intensity for Google Maps and registry food outlet distributions across five validation regions and five outlet categories. NNI values were below 1.0 in nearly all regions and categories for both datasets, indicating consistent spatial clustering in both Google Maps and registry data relative to complete spatial randomness. Agreement between datasets was strongest in the United Kingdom and the Netherlands, where NNI values were closely matched across all outlet categories. The most notable divergences occurred in Denmark, where fast-food outlets showed near-random spatial distribution in the registry (NNI: 0.797) compared to strong clustering in Google Maps (NNI: 0.241), and in Brno, where bars and pubs showed near- dispersed distribution in Google Maps (NNI: 1.353) against strong clustering in the registry (NNI: 0.414), likely reflecting differences in data acquisition timing and registry scope rather than genuine spatial differences between datasets.

**Figure 3.**
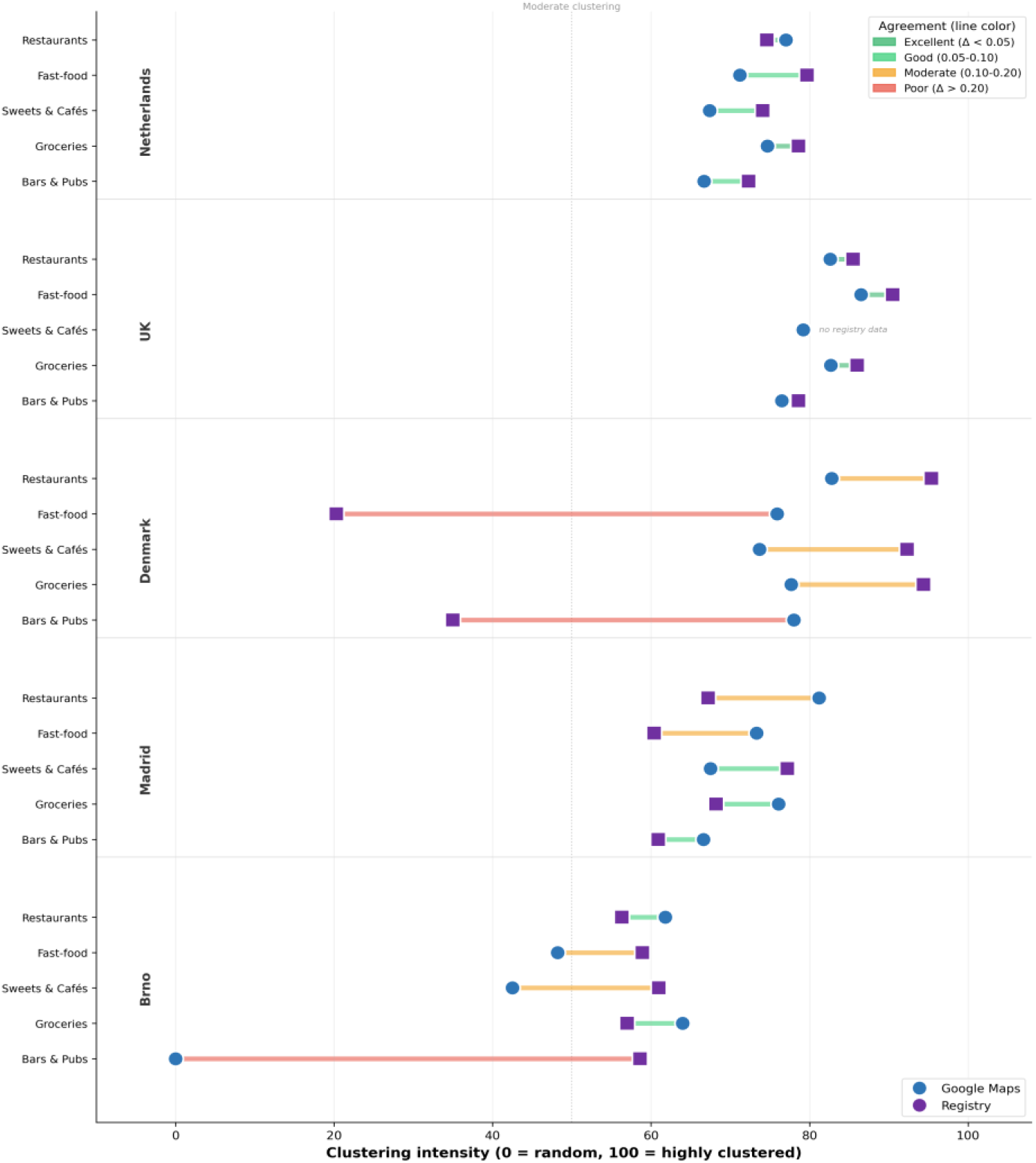
Spatial clustering of food outlets by category and validation region: Google-derived POIs compared to registry data using the Nearest Neighbour Index

Figure 4 presents kernel density estimation (KDE) maps comparing the spatial density of Google - derived food outlet POIs and registry outlets across the five validation regions, along with difference maps. Spatial agreement was high across all cities, with Pearson correlations ranging from 0.94 in Madrid to 0.99 in Copenhagen, Amsterdam, and Brno, indicating strong correspondence in the geographic distribution of food outlet density. London and Madrid showed slightly lower but still strong agreement (r = 0.98 and r = 0.94 respectively), with both cities recording higher registry KDE values than Google Maps, indicating that official registries captured somewhat higher outlet density concentrations in those cities than Google Maps.

**Figure 4.**
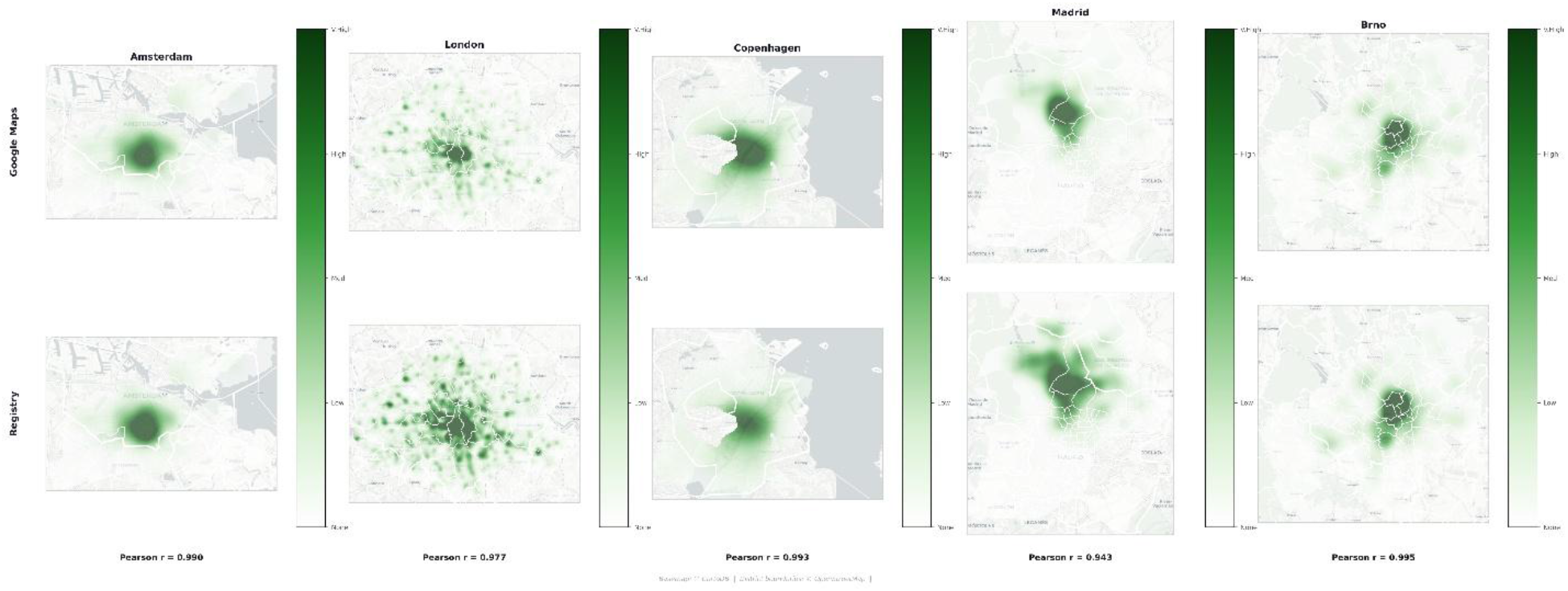
Kernel density estimation (KDE) maps of food outlet spatial density for Google POI and registry datasets across five validation cities with difference maps

### Food outlet composition and distribution across Europe

Figure 5a presents the share of food outlet categories across all study countries. Restaurants constituted the largest category in the majority of countries, with shares exceeding 50% in Italy, Switzerland, Portugal, Luxembourg, Sweden, and Finland. In contrast, groceries and food retail dominated in several Eastern and Southeastern European countries, including Romania, Czech Republic, Poland, Hungary, Bulgaria, and Croatia, where grocery shares ranged from approximately 38% to 49%. Fast-food and snack outlets were most prominent in Bosnia and Herzegovina, Bulgaria, the United Kingdom, and Ireland, each recording shares above 15%, while Italy, Portugal, Austria, and Switzerland showed comparatively low fast-food outlet representation. Sweets and cafés displayed notable variation, exceeding 20% in Greece, Croatia, Latvia, Ireland, and several Baltic and Balkan states. Bars and pubs remained a minor category in most countries but reached double-digit proportions in the United Kingdom, Denmark, the Netherlands, and Liechtenstein.

**Figure 5.**
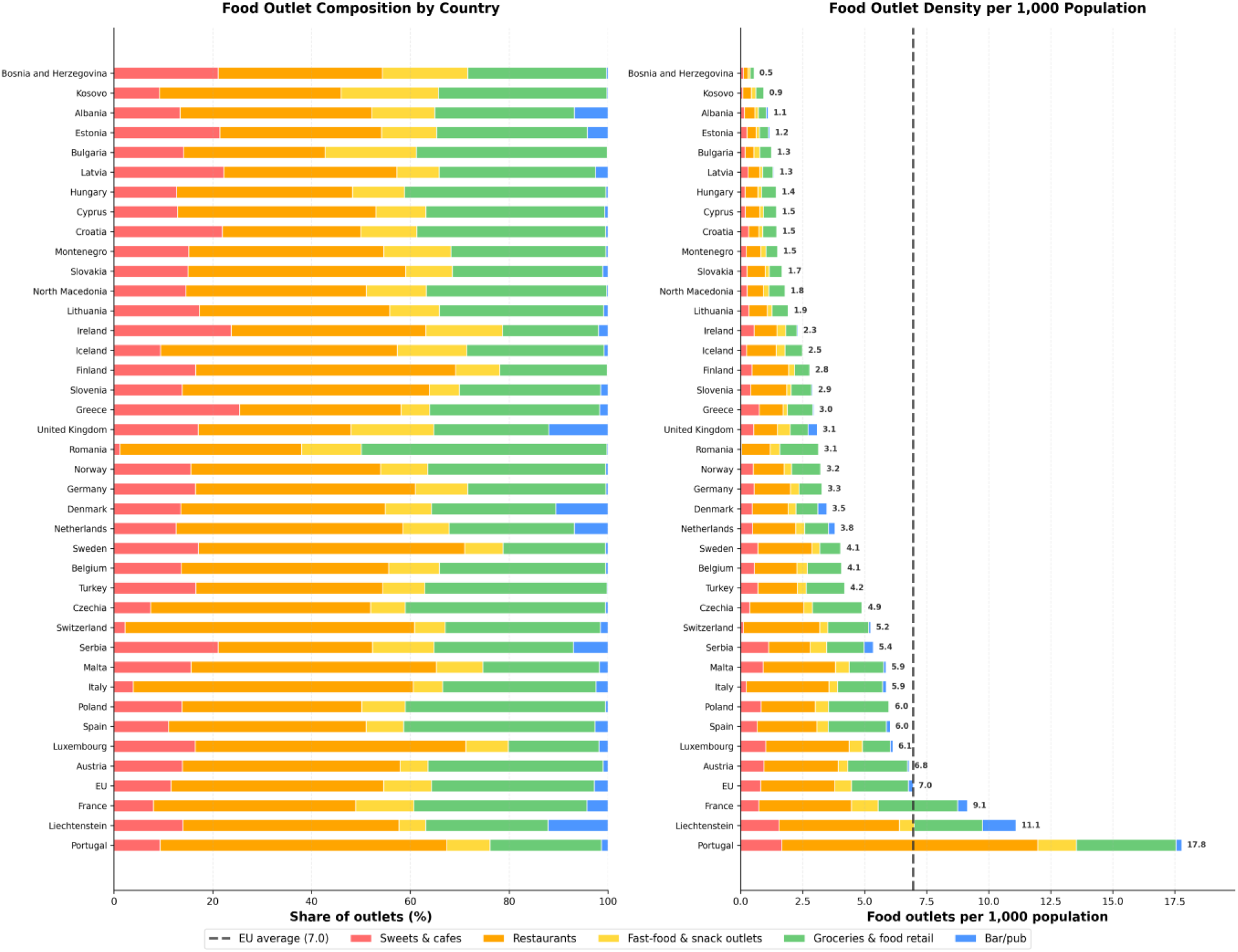
a (left). Share of food outlet categories by country across Europe. Figure 5b (right). Food outlet density per 1,000 population by country across Europe, with the EU average (7.0) shown as a reference line

Figure 5b presents total food outlet density per 1,000 population across all study countries, with the EU average marked at 7.0 outlets per 1,000 residents. Portugal recorded the highest density at 17.8 outlets per 1,000 population, substantially exceeding all other countries, followed by Liechtenstein (11.1) and France (9.1). Austria, Spain, Luxembourg, Poland, Italy, and Malta clustered around the EU average (5.9 to 6.8 per 1,000). Eastern and Southeastern European countries recorded the lowest densities, with Bosnia and Herzegovina (0.5), Kosovo (0.9), Albania (1.1), Estonia (1.2), and Bulgaria (1.3) all falling well below 1.5 outlets per 1,000 population. This pattern indicates a clear west-east gradient in food outlet availability relative to population size across Europe.

The map in figure 6 shows total food outlet density across Europe in z-score deciles relative to the European country mean. Southern and western European countries, particularly Portugal (z = 4.11), France (z = 1.97), and Austria (z = 0.82), fell in the highest deciles, while southeastern and eastern countries such as Bosnia and Herzegovina (z = -1.09), Kosovo (z = - 0.93), and Bulgaria (z = -0.86) consistently occupied the lowest deciles, reflecting a clear west-south to east gradient in food outlet availability.

**Figure 6.**
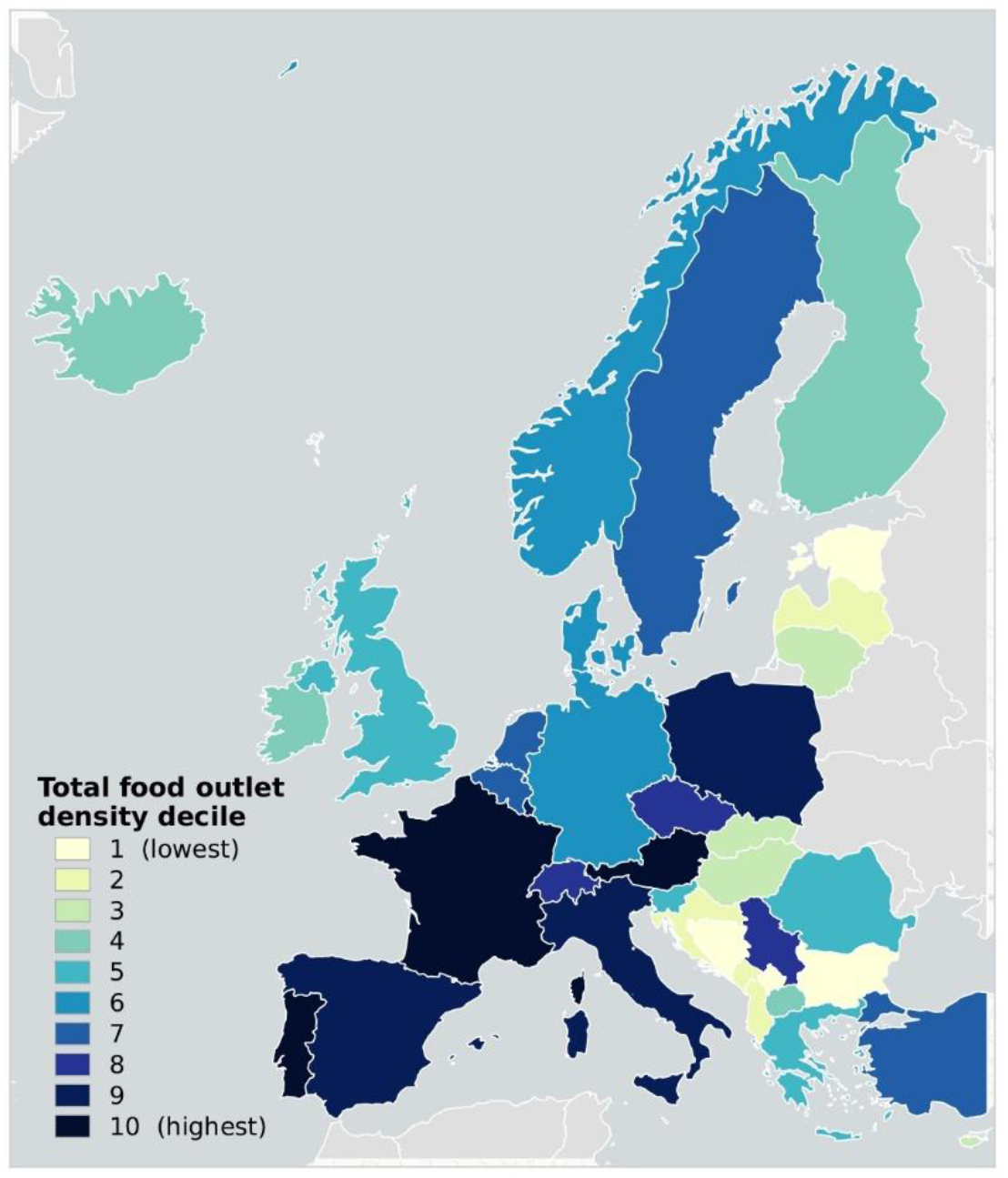
Total food outlet density across Europe (z-score deciles relative to the European mean)

### Characterising joint food environment exposures

Figure 7 shows model fit criteria based on the BIC for latent class solutions tested from two to seven classes, averaged across all eight cities. The BIC improved substantially between two and four classes, with diminishing returns beyond four classes. A four-class solution was therefore selected as optimal across all cities, consistent with prior food environment typology studies (Hobbs et al., 2018; DeWeese et al., 2018).

**Figure 7.**
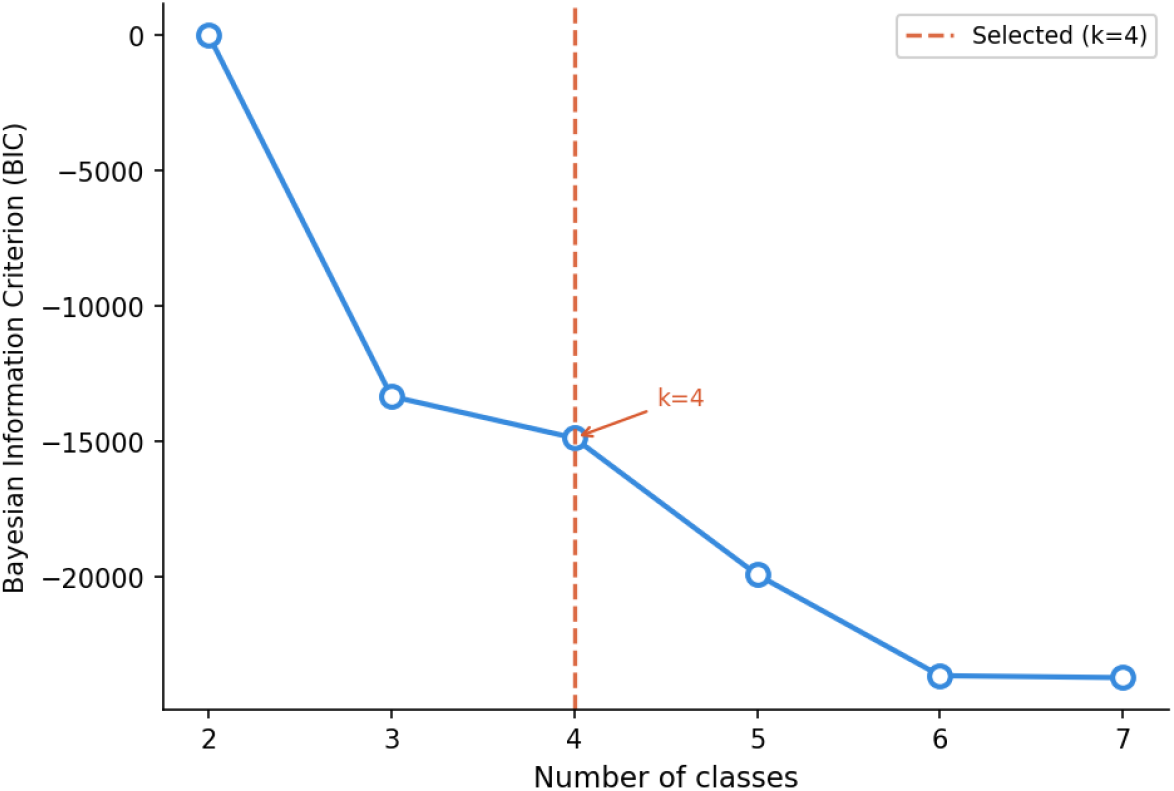
Change in Bayesian Information Criterion (BIC) by number of latent classes across eight European cities

The mean z-score profiles for the four classes are presented in supplementary table S4. Four neighbourhood food environment classes were identified. The ‘low-access’ class was characterised by below-average densities across all five outlet categories simultaneously, reflecting neighbourhoods with limited densities of food outlets of any type. The ‘moderate mixed’ class showed broadly above-average densities across all categories, representing neighbourhoods with moderately active food retail across outlet types. The ‘average access’ class was characterised by outlet densities close to the city mean across all five categories, representing neighbourhoods with typical food outlet exposure without strong specialisation toward any outlet type. The ‘high density mixed’ class displayed strongly elevated z-scores across all five outlet categories simultaneously, with bars and pubs showing the strongest elevation (mean z-score: +3.83), reflecting the concentration of nightlife and hospitality districts within urban commercial cores.

The spatial class distribution varied substantially across the eight cities (Figure 8 and 9). ‘Low-access’ was the most prevalent class across all cities, ranging from 26.0% of hexagons in Paris to 74.7% in Budapest, indicating that the majority of populated urban neighbourhoods are characterised by below-average food outlet density across all outlet types simultaneously. High density mixed was the least prevalent class across all cities, ranging from 1.2% to 16.4%, reflecting the spatially concentrated nature of the most food-dense environments. Western European cities showed higher proportions of Average access and High density mixed hexagons, while Central and Eastern European cities were more strongly dominated by Low-access hexagons, suggesting greater spatial concentration of food retail in Western European urban contexts.

**Figure 8.**
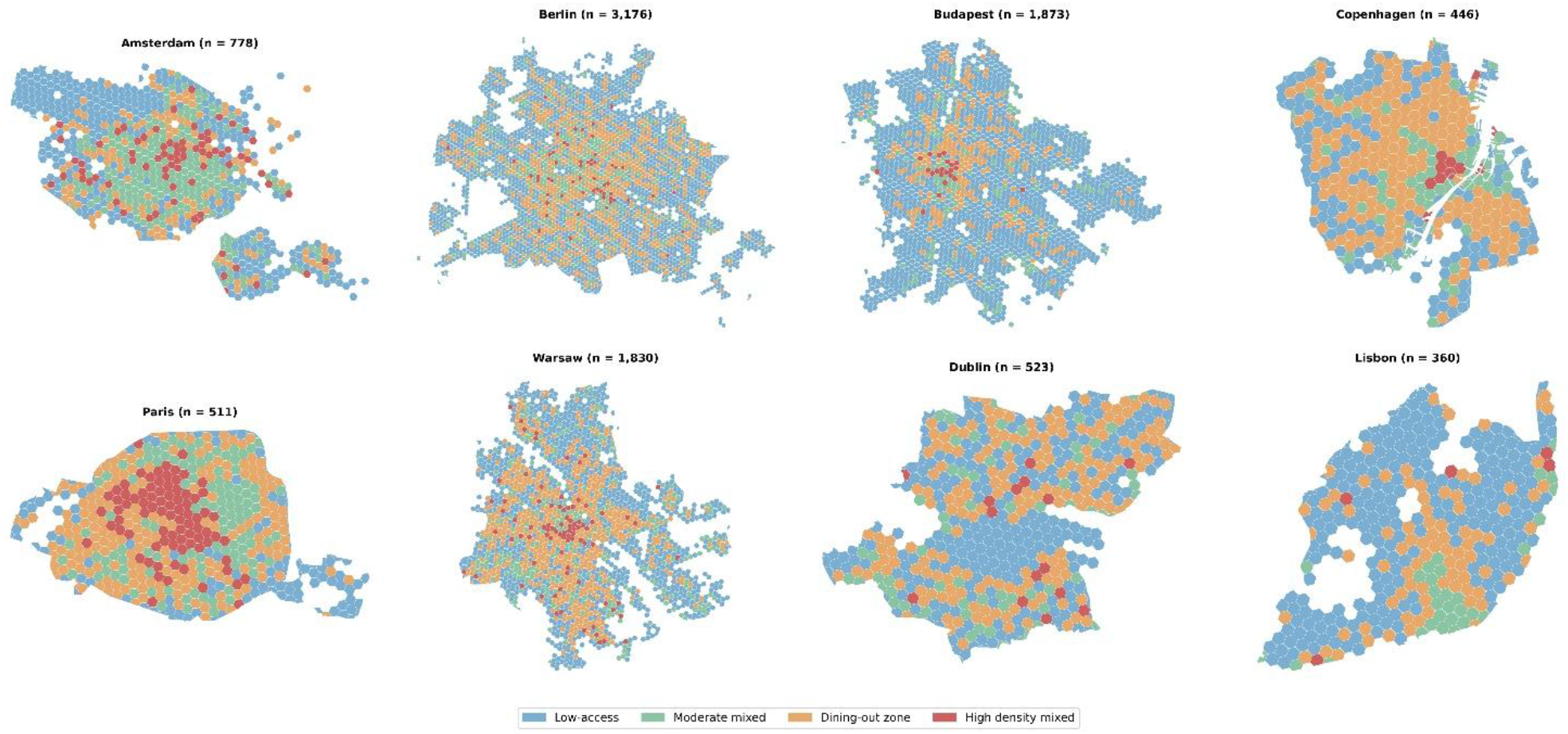
Neighbourhood food environment classes across eight European cities (500m hexagonal grids)

**Figure 9.**
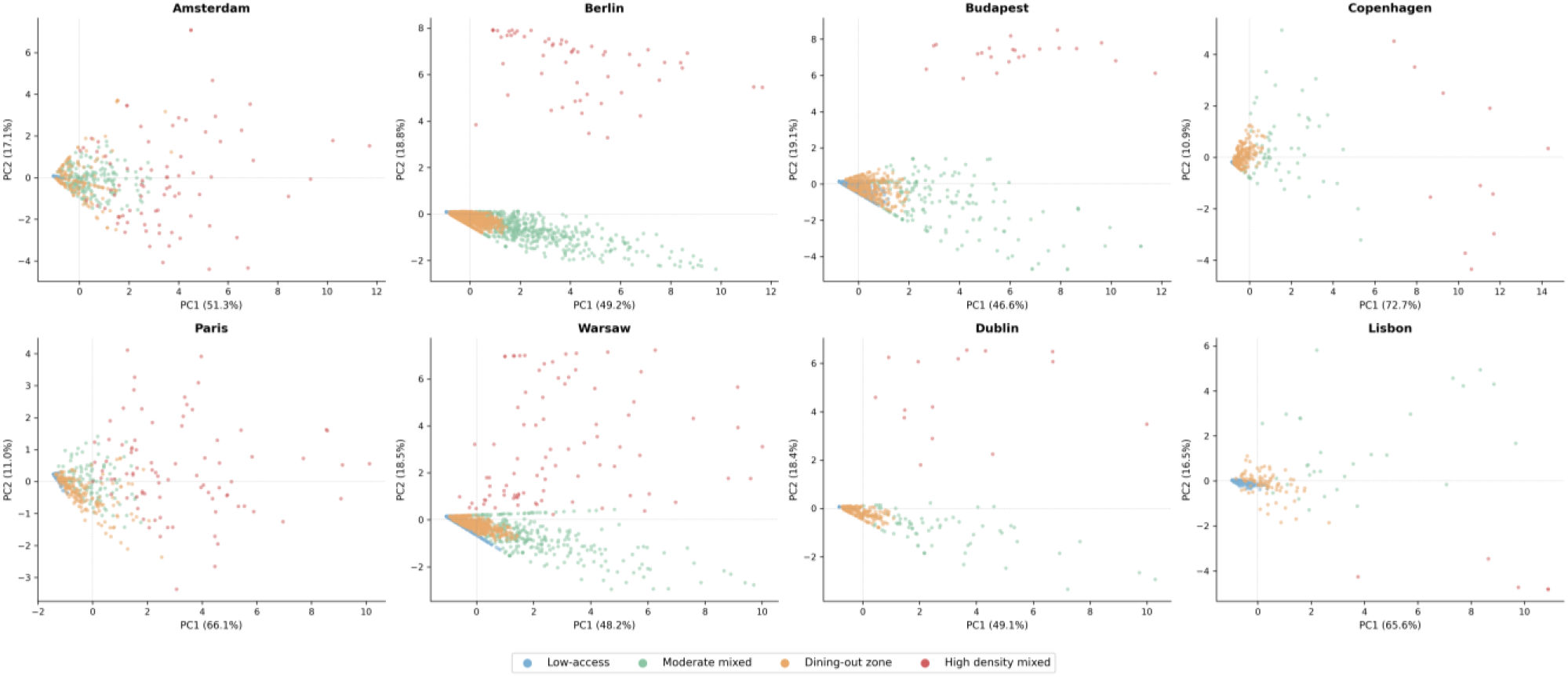
Principal Component Analysis (PCA) scores by neighbourhood food environment class across eight European cities. Each point represents a 500 m hexagon. PC1 reflects overall food outlet density; PC2 reflects a contrast between café/bar and fast-food/supermarket densities. Colours indicate LCA class assignment

## Discussion

This study demonstrates that Google-derived POI data, compiled through a systematic query-based approach, can support high-resolution, cross-national mapping of retail food environments across Europe. Positional accuracy was sufficient for neighbourhood-level spatial analyses across the five validation regions, with median distance errors below 12 m and all matched outlets falling within 30 m of their registry counterpart. These results are comparable to accuracy levels reported in single-city validation studies in Brazil and Mexico (de Menezes *et al*., 2021). Relative completeness was more variable, ranging from 57.4% to 92.2% for registry-to-Google Maps alignment, likely reflecting regional differences in registry scope and temporal currency rather than systematic gaps in Google Maps coverage. Spatial patterning of outlets was consistent between Google Maps and registry datasets across all regions, with KDE Pearson correlations of 0.94 to 0.99, supporting the use of Google Maps as a reliable proxy for the geographic distribution of food retail. At the European scale, food outlet density revealed a pronounced west-east gradient (0.5 to 17.8 outlets per 1,000 population), and co-exposure profiling identified four spatially consistent neighbourhood food environment classes across eight cities, suggesting that urban food environments across Europe share structural patterns despite substantial variation in overall outlet density.

The pronounced west-east gradient in food outlet density observed across Europe, ranging from 0.5 outlets per 1,000 population in Bosnia and Herzegovina to 17.8 in Portugal, reflects broader structural differences in urban development, economic conditions, culture, and food retail market maturity across the continent. Western and Southern European countries, particularly Portugal, France, and Spain, show substantially higher densities driven primarily by restaurants and fast-food outlets, consistent with the well-documented role of eating-out culture and tourism-driven food retail in these contexts (Caspi *et al*., 2012; Wilkins *et al*., 2017). In contrast, Eastern and Southeastern European countries show more grocery-dominated food environments with lower overall densities, which may reflect a greater reliance on home-prepared food and less developed commercial food retail infrastructure. These compositional differences matter for health research: the dominance of fast-food and restaurant outlets in high-density Western European cities has been linked to higher obesity risk in prior meta-analyses (Pineda *et al*., 2024), while the relative scarcity of food retail in Eastern Europe may reflect a different but equally important exposure pattern relating to food access rather than food over abundance (Meijer et al., 2023).

The co-exposure profiling results demonstrate that neighbourhood food environments across European cities share a consistent underlying structure despite substantial variation in overall outlet density. The four-class solution identified through LCA, ranging from Low-access to High density mixed neighbourhoods was replicated consistently across all eight cities, suggesting that the typology captures fundamental patterns of food retail co-location rather than city-specific artefacts. This is consistent with prior typology studies in single-city contexts, where latent class and cluster approaches have similarly identified a small number of interpretable neighbourhood food environment profiles. The PCA findings reinforce this interpretation: the dominant first component, explaining 46.6% to 72.7% of variance across cities, showed broadly equal positive loadings across all five outlet categories, indicating that food outlet types co-occur spatially rather than substitute one another. This co-occurrence pattern has important implications for exposure assessment in epidemiological studies, as it suggests that residents in high-density neighbourhoods are simultaneously exposed to multiple outlet types rather than to any single category in isolation, complicating efforts to attribute health outcomes to specific food environment dimensions (Pinho et al., 2019).

### Strengths

A key strength of this study is the combination of broad geographic coverage and high spatial resolution. By compiling and validating food-related POIs across 39 countries in the European study region, and constructing exposure measures at 500 meter grid-cell resolution, this study provides a level of cross-national comparability that has rarely been available for food environment research in Europe. The harmonised classification system, comprising five main categories and twenty five sub-categories, was developed to accommodate over 600 unique Google Places type variants, enables more consistent comparisons across countries with different food retail terminology, business registration systems, and retail structures.

Another strength is the use of a multi-metric validation framework. By combining positional accuracy, bidirectional relative completeness, NNI, and KDE, we assessed several complementary dimensions of data quality than single-metric approaches common in prior studies (Bader et al., 2010; Zhao et al., 2020), and the use of five geographically and institutionally diverse validation regions strengthens the generalisability of the validation findings. The co-exposure profiling approach, combining PCA and LCA across eight cities, demonstrates that the dataset supports methodologically sophisticated exposure characterisation beyond simple outlet counts or density measures. Finally, by making all code openly available, reproducible and scalable framework for harmonised food environment exposure assessment in support of urban health, spatial epidemiological, and comparative place-based research.

### Limitations

Several limitations must be acknowledged. First, no gold standard reference dataset was available for any validation region; official registries used as reference sources are themselves subject to incompleteness, temporal mismatch, geocoding errors, and classification inconsistencies, which likely contributed to the variability in bidirectional completeness observed across regions. Second, temporal misalignment between Google-derived POI data, which reflects near continuously updated from user and merchant contributions, and registry datasets, which are typically updated annually or biannually, introduces systematic discrepancies that cannot be fully disentangled from genuine coverage gaps (Pinho et al., 2019). Third, classification heterogeneity across countries complicates cross-national comparisons; while the harmonised classification reduces inconsistency, local variation in how businesses register and describe themselves means that some misclassification is unavoidable, particularly for subcategories such as bakeries, dessert shops, and convenience stores (Liese et al., 2022). Fourth, the validation was conducted in five regions concentrated in Western and Central Europe, limiting conclusions about data quality in Eastern and Southeastern European countries where registry systems are less comprehensive and Google Maps coverage may be less complete. Future studies should prioritise ground-truthing through field audits or Street View verification in these underrepresented regions, and incorporate temporal tracking of POI changes across sequential snapshots to better quantify update lags between Google Maps and official data sources.

Beyond methodological contributions, this work provides the first harmonized, Europe-wide representation of the neighborhood food environment, enabling researchers to link spatial food outlet patterns to dietary behaviour and health outcomes across countries.

## Conclusions

This study establishes a methodological foundation for systematic, high-resolution, cross-national food environment research across the European study region. We show that Google-derived POI data, when compiled through a structured and reproducible query strategy, harmonised through a standardised classification system, and validated against registry datasets, can reliably characterise key spatial patterns in the retail food environment at continental scale. The openly available code, data (available upon request) and harmonised classification system developed in this study provide a scalable resource for food environment monitoring, comparative spatial analysis and other research studies including epidemiological analyses. This methodological framework may be transferable to other urban exposure domains, supporting efforts to characterise, compare, and monitor urban exposures across diverse geographic contexts.

## Data Availability

All data produced in the present study are available upon reasonable request to the authors

https://github.com/mohanrajujs/OBCT.git

## Data availability

The GitHub link has been included here: https://github.com/mohanrajujs/OBCT.git

